# Double COVID-19 Confirmed Case Fatality Rate in Countries with High Elderly Female Vitamin D Deficiency Prevalence

**DOI:** 10.1101/2020.06.13.20130484

**Authors:** Alex Bäcker, Myron Mageswaran

## Abstract

A number of clues point to a possible role of vitamin D in fighting COVID-19: a reduction in case growth speed with solar zenith angle, higher fatality rate in black people, lower fatality rate in populations that spend more time outdoors. Yet a direct demonstration that vitamin D deficiency is associated with COVID-19 fatalities has remained elusive. We show here in a comparison of 32 countries that countries with high prevalence of vitamin D deficiency among elderly females show a confirmed case fatality rate twice as high as those with low prevalence. We then show that this effect cannot be explained by differences in life expectancy between countries. A mechanistic role for vitamin D in the severity of COVID-19 is proposed.

**One Sentence Summary:** Vitamin D deficiency among elderly females is associated with countrywide COVID-19 confirmed case fatality rates up to twice as high as those of countries with low vitamin D deficiency prevalence.

## Main Text

The world is in the grip of the COVID-19 pandemic. Public health measures that can reduce the risk of infection and death in addition to quarantines are desperately needed.

The world is undergoing the largest public policy experiment ever conducted. Billions of people are being told to stay at home to avoid COVID-19 contagion. While isolation undoubtedly helps prevent contagion, having people stay at home and closing public parks has side effects that are not fully understood. UV radiation in natural sunlight stimulates vitamin D production in the hours and days following exposure^1^. At least a billion people worldwide are estimated to be vitamin D deficient, mainly because of inadequate exposure to sunlight and inadequate fortification of food with vitamin D^2^. Interventional and observational epidemiological studies provide evidence that vitamin D deficiency may confer increased risk of influenza and respiratory tract infection^3,4^. Vitamin D deficiency is also prevalent among patients infected with another RNA virus, HIV. Cell culture experiments support the thesis that vitamin D has direct anti-viral effects particularly against enveloped viruses. Though vitamin D’s anti-viral mechanism has not been fully established, it may be linked to vitamin D’s ability to up-regulate the anti-microbial peptides LL-37 and human beta defensin 2^5^. Sunlight exposure improves survival in patients for some diseases^6^. The SARS coronavirus is inactivated under ultraviolet irradiation^7^. Viruses containing single-stranded nucleic acids such as coronaviruses are more sensitive, unable to repair damage in the absence of a complementary strand. Vitamin D deficiency has been associated with increased risk of autoimmune diseases, infectious diseases and cardiovascular disease^8^. Studies from the Netherlands and France suggest that clots arise in 20–30% of critically ill COVID-19 patients9,10. Vitamin D has anticoagulant effects^11,12^.

Are governments inadvertently putting populations at risk by locking them away from the sunlight for months at a time? Should public policy to slow the COVID-19 pandemic include both social isolation and sun exposure or vitamin D intake?

Portugal, which borders Spain, has a COVID-19 mortality rate 75% lower than Spain. Norway and Denmark have low numbers of COVID-19 deaths, too, despite low winter sunlight levels. Addition of vitamin D to margarine is compulsory in Norway, Denmark, the Netherlands, Belgium and Portugal.

COVID 19 is much more lethal to the elderly, and older populations tend to have much higher levels of vitamin D insufficiency^13,14,15^.

43% of US COVID 19 deaths have occurred in nursing homes^16^. 84-93% of US nursing home patients have insufficient vitamin D levels^17,18^.

Relevant amounts of vitamin D are found in seafood, egg yolk, butter and some mushrooms. Japan, a country with high fish consumption^19^, has been relatively spared, with less than 3% of the COVID 19 mortality of the US^20^.

The synthesis of vitamin D in human skin is a two-stage process that begins with the production of previtamin D after irradiation of 7-dehydrocholesterol by ultraviolet (UV) radiation. A number of factors control the probability of a suitable UV photon reaching a molecule of 7-dehydrocholesterol in the skin. These are astronomical factors that govern the solar zenith angle (SZA), and the local state of the atmosphere, determining the available solar UV radiation; skin pigmentation and age, determining competing absorbers of UV radiation and available 7-dehydrocholesterol; individual behavior in the local surroundings, determining exposure of unprotected skin to available UV radiation. The only one of these influences that can be determined unequivocally for any situation is the SZA. At noon, SZA is at a minimum and is equal to latitude minus solar declination angle. At large SZAs there is insufficient solar UV radiation to initiate significant vitamin D synthesis^21^.

One of us has previously shown SZA is the best predictor we know for COVID-19 case growth speed^22^.

And yet, the topic remains controversial, and recent reviews maintain there is no evidence of a link between vitamin D and COVID-19^23,24,25,26^.

To address this question, we compared the prevalence of severe vitamin D deficiency (25-hydroxyvitamin D serum concentration below 12 ng/ ml (30 nmol/l)) and intake levels in different countries^27,28,29,30,31,32,33,34^, with COVID-19 case growth rates and confirmed case fatality rates (cCFR).

COVID-19 case growth speed was not significantly correlated with the prevalence of subclinical vitamin D deficiency in the female elderly population. This result is consistent with a recent study of the UK Biobank^35^.

A multiple linear regression model showed a significant correlation between both vitamin D deficiency in the female, but not male, elderly population and cCFR (Fig.1, p<0.05). The significant association with vitamin D deficiency prevalence is particularly notable given that the seasonality of vitamin D levels is likely to have introduced noise in the data. Furthermore, the fact that the effect is significant even with national averages, which mask the effect in individuals who are vitamin D deficient with others who are not, suggests the effect is likely much stronger when comparing individuals with different vitamin D levels.

**Figure 1.**
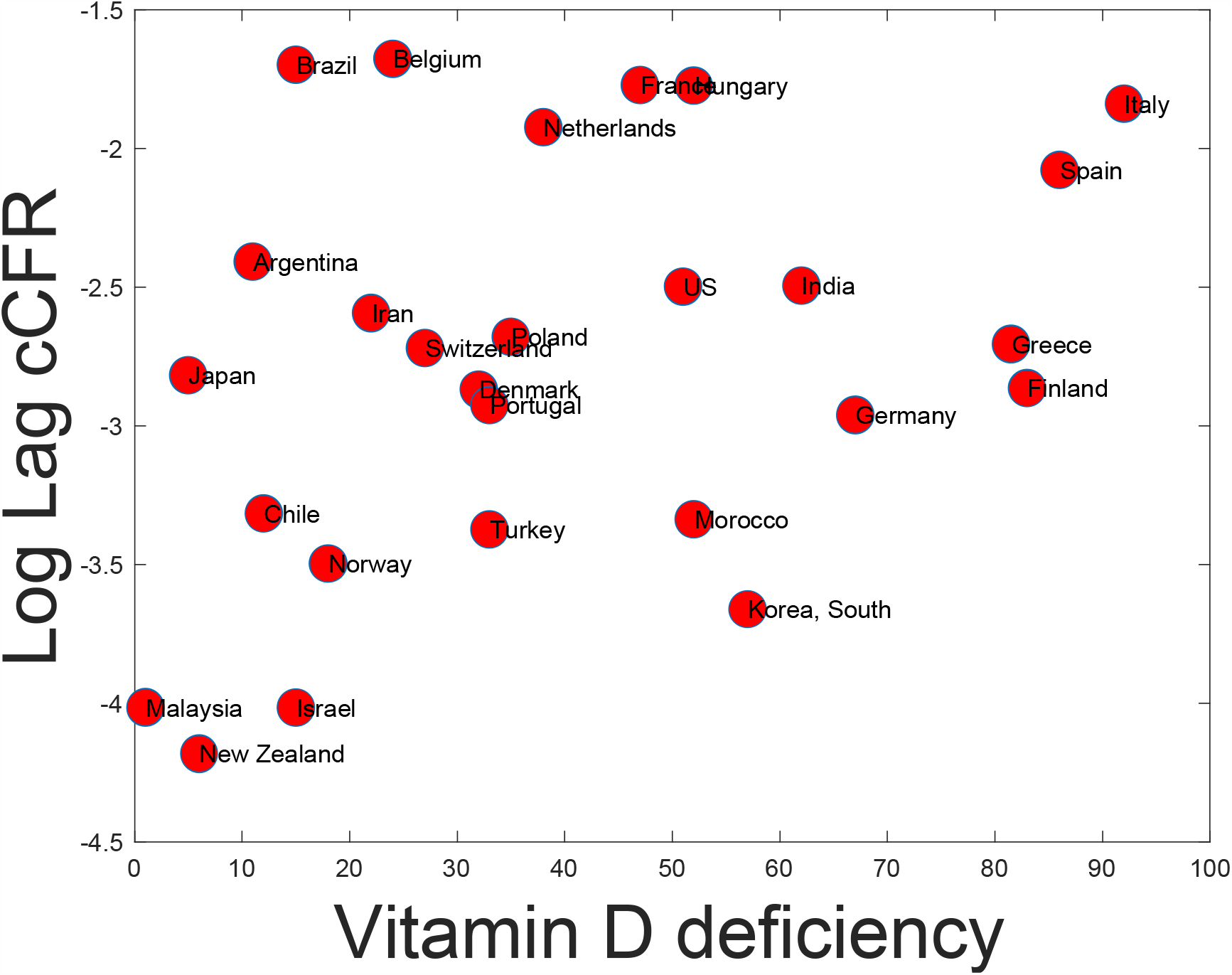
cCFR is significantly higher in countries with high prevalence of vitamin D deficiency among elderly females (p<0.05).

Noticing that some of the countries with high cCFR yet low vitamin D deficiency, such as Brazil, had a large black population, and given our previous result showing COVID-19 mortality is higher in Black populations in low irradiance locations^63^, we carried out the same analysis correlating COVID-19 cCFR with the percentage of the population that is Black, and found a significant positive correlation (Fig. 2, p<0.05).

**Figure 2.**
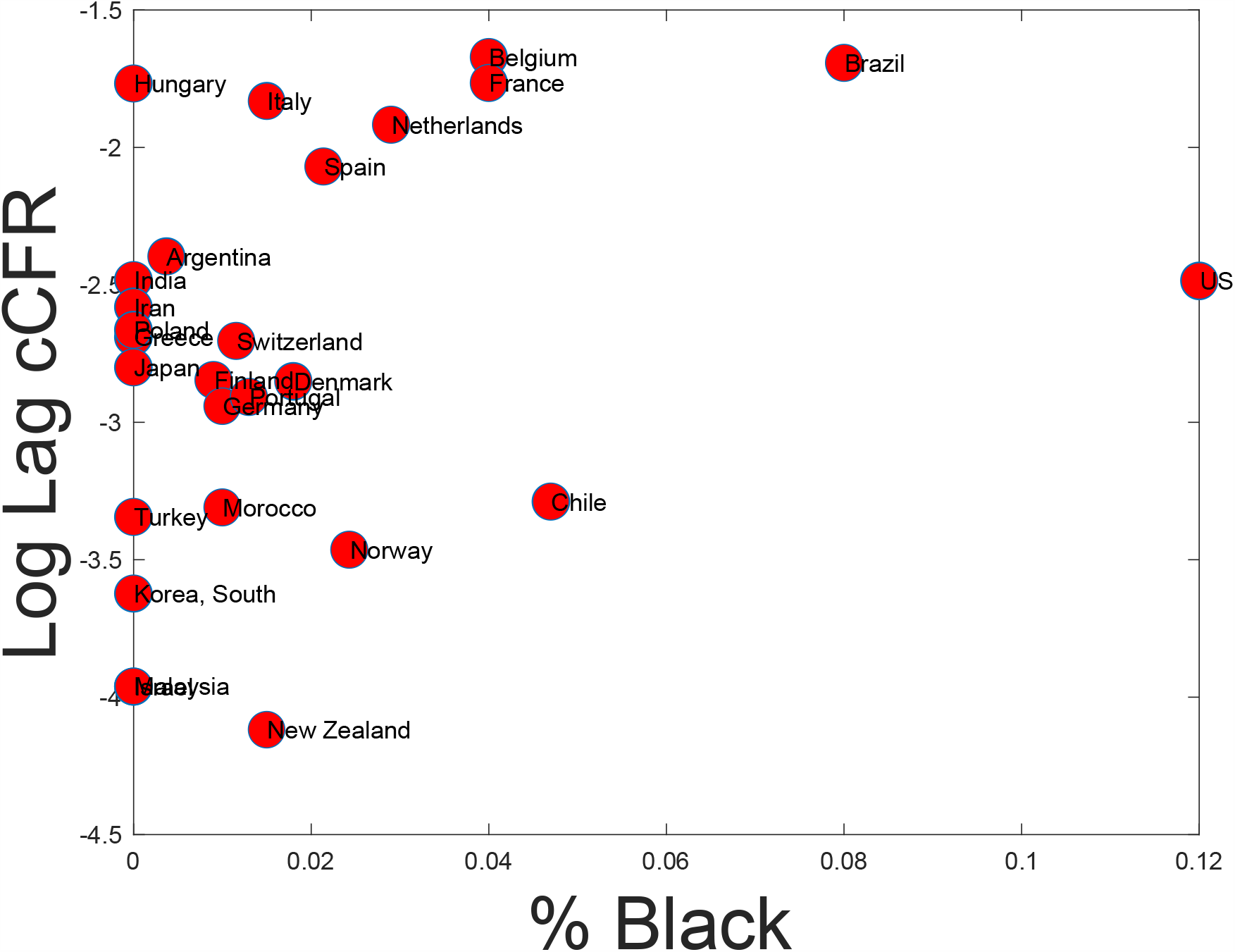
cCFR is significantly higher in countries with high percentage of black inhabitants (p<0.05).

To disambiguate the impact of each of these effects, we developed a multivariate linear model to estimate the significance of vitamin D deficiency prevalence and black population % when evaluated jointly. Both factors were significantly associated with COVID-19 cCFR (Table 1).

**Table 1.** Impact of vitamin D deficiency prevalence among elderly women and the percentage of the population that is Black on COVID-19 cCFR:

|  | Coefficient | SE | p-Value |
| --- | --- | --- | --- |
|  | _____ | _____ | _____ |
| (Intercept) | -3.3044 | 0.23164 | 3.2241e-13 |
| % population Black: | 10.585 | 4.4673 | **0.026205** |
| Vitamin D deficiency: | 0.010317 | 0.0047414 | **0.039629** |

Number of observations: 27, Error degrees of freedom: 24
Root Mean Squared Error: 0.645
R-squared: 0.296, Adjusted R-Squared: 0.237
F-statistic vs. constant model: 5.04, p-value = 0.0148

The prevalence of vitamin D deficiency among elderly females was not significantly correlated with the % of Blacks in the population.

Vitamin D intake was not significantly correlated with case growth rate or with cCFR.

The elderly are particularly susceptible to COVID-19. Even in low latitudes where severe vitamin D deficiency is rare, there is frequent occurrence of subclinical vitamin D deficiency, especially in elderly people^36^.

Kumar and Srivastava recently claimed that average life expectancy could explain the apparent impact of vitamin D deficiency prevalence on COVID 19^37^. However, that study calculated cCFR with no lag between deaths and cases, which produces an inaccurate estimate of fatality. It also used a relatively small number of countries. We addressed both of those issues. We estimated confirmed case fatality rate as the average ratio between the number of new observed deaths each day and the number of new deaths that would have been expected given the number of new cases per day convolved by the diagnosis to death lag curve, as described in the literature^38^. A linear model incorporating all three factors showed vitamin D deficiency prevalence, but not life expectancy^39^ or percentage of the population black, were positively and significantly correlated with COVID-19 cCFR (Table 2; p<0.05). Interestingly, we were able to replicate Kumar and Srivastava’s result when cCFR was measured the way they did, with constant zero lag between cases and deaths. Yet such an estimate fails to account for the significantly lower of cases that existed one lag before in an exponentially growing epidemic, and/or the number of cases yet to die. There was no significant correlation between each country’s cCFR and life expectancy, or between life expectancy and vitamin D deficiency prevalence.

**Table 2:** Impact of the percentage of the population that is Black, vitamin D deficiency prevalence among elderly women and life expectancy on COVID-19 cCFR:

|  | Estimate | SE | tStat | pValue |
| --- | --- | --- | --- | --- |
|  | _____ | _____ | _____ | _____ |
| (Intercept) | 5.5474 | 6.6922 | 0.82895 | 0.41646 |
| % population Black: | 14.282 | 10.311 | 1.3851 | 0.18056 |
| Vitamin D deficiency: | 0.024229 | 0.010697 | 2.265 | **0.034217** |
| Life expectancy: | -0.098322 | 0.083669 | -1.1751 | 0.2531 |

Number of observations: 25, Error degrees of freedom: 21
Root Mean Squared Error: 1.43
R-squared: 0.27, Adjusted R-Squared: 0.166
F-statistic vs. constant model: 2.6, p-value = 0.0795
Root Mean Squared Error: 0.622
R-squared: 0.295, Adjusted R-Squared: 0.194
F-statistic vs. constant model: 2.92, p-value = 0.0578

## Discussion

A retrospective cohort study of active and expired confirmed COVID-19 cases in Indonesia showed that when controlling for age, sex, and comorbidity, vitamin D status was strongly associated with COVID-19 mortality outcome of cases: vitamin D insufficiency showed an odds ratio (OR) for mortality of 7.63, and vitamin deficiency showed an OR of 10.1240.

In a study performed at a single, tertiary care academic medical center in which the medical records of COVID-19 patients were retrospectively reviewed, the vitamin D insufficiency prevalence in ICU patients was 84.6%, vs. 57.1% in floor patients. Strikingly, 100% of ICU patients less than 75 years old had vitamin D insufficiency^41^.

An analysis of global COVID-19 data using Causal Inference, constructing two contrasting models for vitamin D’s role in COVID-19 fatality, one causal and one acausal, setting out clearly multiple predictions made by each model, showed that observed data strongly matched predictions made by the causal model^42^.

One in two fatal cases of COVID-19 experience a cytokine storm, 82% of whom are over the age of 60^43^. In older individuals, NLRP3 may be poised for hyperactivation by SARS-CoV-2 components^25^. Elimination of severe vitamin D deficiency reduces the risk of high CRP levels (odds ratio of 2) which may be used as a surrogate marker of cytokine storm^44^. Vitamin D inhibits NLRP3 activation by impeding its deubiquitination mediated by BRCC3^45^. Vitamin D downregulates NLRP3 and protects against toxin-induced cell death^46^ and modulates the NLRP3 inflammasome activation^47^. In addition, likely due to its role in vasodilation and reducing inflammation, ACE2 partially protects against sepsis-induced- and SARS-induced severe acute lung injury in mice^48^. Vitamin D can increase the expressions of ACE2 mRNA and protein levels of ACE2 in rats^49^. Children have higher ACE expression than adults^50^, which could help explain their relative immunity to COVID-19. ACE2 expression is dramatically reduced with aging in rats of both genders: young-adult vs. old *P* < 0.001 (by 78% in male and 67% in female, respectively) and middle-aged vs. old *P* < 0.001 (by 71% in male rats and 59% in female rats, respectively). The decrease of ACE2 content is relatively slight between young-adult and middle-aged groups (by 25% in male and 18% in female, respectively). Although there is no gender-related difference of ACE2 in young-adult and middle-aged groups, a significantly higher ACE2 content was detected in old female rats than male. In conclusion, the more elevated ACE2 in young adults as compared to aged groups may contribute to the predominance in SARS and COVID-19 attacks in aged populations^51^.

While men and women have the same COVID-19 prevalence, men with COVID-19 are more at risk for worse outcomes and death, independent of age^52^. Estrogen and testosterone impact immune system response and engagement, resulting in a less robust immunologic response in males and subsequent increased morbidity and mortality from viral respiratory illnesses^53^. Several animal models have demonstrated increased ACE2 activity in the male or ovariectomized model^54^.

Males show an upregulation of angiotensin II receptor type 1 (AGTR1), a member of the renin-angiotensin system (RAS) that plays a role in angiotensin-converting enzyme 2 (ACE2) activity modulation. *In vivo* studies have showed that ovariectomized females, in addition to males, usually have higher levels of ACE2 activity compared to non-ovariectomized females. This might indicate a possible role of the sex hormones in regulating ACE2 activity. Many reports highlight the role of human receptor ACE2, which also belongs to the renin-angiotensin system (RAS), in facilitating the binding of the SARS-CoV-2 to the host cells. While AGTR2 showed no significant difference in its expression between both genders, AGTR1 expression levels were higher in tissues obtained from male individuals compared to females^55^.

Female gender is associated with lower vitamin D levels (14.5 ± 10.9 vs. 15.9 ± 9.5, *p* = 0.007) and independently associated with severe vitamin D deficiency (41.9% vs. 30.4%, *p* < 0.001; adjusted odds ratio (OR) (95% confidence interval (CI)) = 1.42 (1.08–1.87), *p* = 0.01). Yet note that a French and Canadian study reported elderly males suffered from a serious vitamin D deficiency as compared to elderly females^56^.

Vitamin D and vitamin D receptor agonists modulate the immune system against over-reactivity towards tolerance; on this basis, vitamin D has been proposed as a therapeutic candidate to control autoimmune processes. Females retain greater immune reactivity and competence likely due to estrogens, which, at variance with androgens, are associated with a greater resilience to infections but also to a higher risk for autoimmunity^57^. Vitamin D deficiency is considered a risk factor for autoimmune diseases. Vitamin D and its analogues have been proposed as therapeutic tools in autoimmunity considering their exquisite immunoregulatory effect against over-reactivity towards tolerance. Beneficial effects against autoimmune processes seem to be allowed by vitamin D acting in synergy with estrogens^58^.

These facts may explain why it’s vitamin D deficiency among older females that is the stronger predictor of COVID-19 cCFR.

DeGroot and Bontrop^59^ have pointed out that one of the prominent protein families that senses infection, and that is essential to the innate immune system, comprises the Toll-like receptors (TLR) (Akira and Takeda 2004): In humans, there are 10 different members, and each of them can be activated by different types of ligands stemming from disparate pathogens. TLR play a role in clearing infections. The TLR types present on intracellular membranes of endosomes (TLR3, 7, 8, and 9) are intended to register, though not exclusively, viral infections by detecting foreign types of nucleic acids (Fitzgerald and Kagan 2020). The nuclear material of coronaviruses is single-stranded RNA (ssRNA). The structurally related TLR7 and 8 tandem is encoded by the female X chromosome (Armant and Fenton 2002). Therefore, these latter two genes may represent gender-related risk factors. The prominent ligand for TLR7 and 8 is viral ssRNA (Diebold et al. 2004; Heil et al. 2004) and, more specifically for TLR8, its RNAse T2 degradation products (Greulich et al. 2019). In humans, TLR7 is constitutively expressed on the membranes of endosomes in plasmacytoid dendritic cells (DC) and B lymphocytes, whereas

TLR8 is more prominently present in cells of the myeloid lineage such as monocytes and neutrophils (Hornung et al. 2002). TLR7-positive plamacytoid cells are copiously present in lung tissue (Plantinga et al. 2010). Upon activation of the TLR7 pathway, for instance by the influenza virus, plasmacytoid DC have the capacity to produce significant amounts of type I interferon (Di Domizio et al. 2009). Moreover, TLR8 is present in the lung, and is known to act upon infection by various viruses (Beignon et al. 2005; Heil et al. 2004). An immune response activated via the TLR7 and/ or 8 pathway may also induce unwanted side effects. SARS-Cov-1, for instance, may induce a cytokine storm (Tang et al. 2016), and the viral activation of neutrophils in the case of asthma may result in lung pathology (Li et al. 2013). One of the two female sex chromosomes is generally inactivated in females (Lyon 1992; Schurz et al. 2019). In female immune cells, however, TLR7 genes – and probably TLR8 – escape such silencing. As a consequence, the genetic information on both chromosomes is expressed, whereas male individuals possess only a single copy of the X chromosome (Souyris et al. 2018). Gender-related expression profiles may result in a prominent dosage effect in which females are more prone to activate an immune response to single-stranded viruses such as SARS-CoV-2.

Whilst males may be more vulnerable to the respiratory symptoms of COVID-19, women may be more likely to show the auto-immune like inflammatory response that appears to be modulated by vitamin D.

Vitamin D deficiency and insufficiency occurs in 29.2% and 54.1% of black women, respectively, contrasting with 5% and 42.1% of white women^60^.

A study of 660 women in South Los Angeles showed the prevalence of vitamin D deficiency among Hispanic women was 45% lower than among African American women^61^. One of us has previously shown that non-Black Hispanics have a significantly lower COVID-19 cCFR than non-Hispanics^64^.

Any benefit of vitamin D needs to be balanced against the risk of toxicity, which is characterized by hypercalcemia. Daily brief, suberythemal exposure of a substantial area of the skin to ultraviolet light, climate allowing, provides adults with a safe, physiologic amount of vitamin D, equivalent to an oral intake of about 10,000 IU vitamin D_3_ per day, with the plasma 25-hydroxyvitamin D (25(OH)D) concentration potentially reaching 220 nmol/L (88 ng/mL). The incremental consumption of 40 IU/d of vitamin D_3_ raises plasma 25(OH)D by about 1 nmol/L (0.4 ng/mL). High doses of vitamin D may cause hypercalcemia once the 25(OH)D concentration is well above the top of the physiologic range. Evidence from clinical trials shows, with a wide margin of confidence, that a prolonged intake of 10,000 IU/d of vitamin D_3_ poses no risk of adverse effects for adults, even if this is added to a rather high physiologic background level of vitamin D^62^.

Inaction can be just as dangerous as action in the face of uncertainty. Several converging lines of evidence now suggest a role for sunlight in the prevention and treatment of COVID-19: sunnier places have slower COVID-19 growth^22^, African Americans and Black people in the UK have higher COVID-19 fatality rates in low irradiance locations^63^, US Latinos have higher sunlight exposure and lower COVID-19 fatality rates^64^, vitamin D-deficient populations such as the elderly and those in nursing homes have higher COVID-19 mortality rates, and vitamin D deficiency is associated with significantly higher COVID-19 fatality rates.

Although the possibility remains that another variable correlated with vitamin D (such as sunlight or something correlated with sunlight) is behind the observed effects, we propose that severe COVID-19 shows some characteristics of a virus-induced autoimmune disease, and that both sunlight and vitamin D be tested for its prevention and treatment.

Whilst vitamin D supplementation has been causally linked in a randomized, double-blind, placebo controlled trial to the downregulation of interleukin-1 expression in peripheral blood mononuclear cells in humans^65^, and interleukin-1 Beta (IL-1β) is a critical component of lung inflammation during viral infection^66^, it should be noted that no significant correlation was found between vitamin D intake and COVID-19 fatality. Until a clinical trial of the impact of vitamin D supplementation on COVID-19 is complete, when sunlight is available, the safest course of action may be for populations vulnerable to COVID-19 may be to simply take a little daily dose of sunshine.

## Methods

Vitamin D levels are season-dependent. In Chile, for example, 20% of post-menopausal women have serum 25OHD concentrations below 15 ng/ml in summer, but 60% of them show that level of deficiency in winter. To address this issue, where possible, we used vitamin D deficiency during the season that the COVID-19 epidemic broke out in each country.

COVID-19 case growth rates were assessed by computing the number of cases on the Nth day after the 100^th^ case in each location. This method aligns by stage in the epidemic, is robust to later government interventions, and to idiosyncrasies about the first few cases in each location.

cCFR were computed by dividing the number of COVID-19 deaths in each country on each day between the start of the COVID-19 pandemic and June 10^th^ 2020 by the convolution of the number of cases in days 42 to 7 before and the distribution of lags between diagnoses and deaths^67^.

## Data Availability

All data are available.

https://docs.google.com/spreadsheets/d/19VyevkN4hp_oNgXJkwMpav7-9Xc7yDfjqt19M3f_SiU/edit?usp=sharing

## Acknowledgments

We are grateful to Prof. Ivan Dmochowski of the University of Pennsylvania and Prof. Armin Schwartzman of UCSD for stimulating discussions, and to Mathworks Inc. for financial support. No competing interests to declare. All data are available at https://tinyurl.com/vitamind4covid19. This paper evolved from earlier work published March 23^rd^ 2020 onwards <u>here</u>, <u>here</u> and <u>here</u>.

